# Risk of Hair Loss with Semaglutide for Weight Loss

**DOI:** 10.1101/2025.02.23.25322568

**Authors:** Mohit Sodhi, Ramin Rezaeianzadeh, Abbas Kezouh, Connor Frey, Mahyar Etminan

**Affiliations:** Epilytics, Vancouver, British Columbia, Canada; Department of Emergency Medicine, Faculty of Medicine, University of British Columbia, Vancouver, Canada; Faculty of Pharmaceutical Sciences, University of British Columbia, Vancouver, Canada; StatExpert Ltd, Laval, Quebec, Canada; MD Undergraduate Program, Faculty of Medicine, University of British Columbia, Vancouver, Canada

**Author notes:** Corresponding author: **Mahyar Etminan**, PharmD, MSc.,Faculty of Medicine | University of British Columbia, Departments of Ophthalmology and Visual Sciences, Pharmacology and Medicine | The Eye Care Center, Room 323-2550 Willow Street, Vancouver BC, V5Z 3N9, |.

**Keywords:** GLP-1 agonist, semaglutide, weight loss, hair loss, drug safety

## Abstract

GLP-1 (glucagon-like peptide-1) agonists including semaglutide are some of the most prescribed medications in the world. Many recent studies have quantified their adverse events with a focus on gastrointestinal adverse events. However, no large epidemiologic study has investigated the risk of hair loss with these medications. We conducted a retrospective cohort study using a large health claims database to investigate this risk. Using a random sample of 16 million patients, we identified new users of semaglutide and an active comparator bupropion-naltrexone. We excluded those with a diagnosis of diabetes or use of antihyperglycemics prior to cohort entry. Cohort members were followed from the first date of a study drug prescription to the diagnosis of hair loss. We adjusted for age, sex, geographic location, depression, steroid use, hypothyroidism, polycystic ovary syndrome, and anemia. Our cohort included 1,926 semaglutide users and 1,348 users of bupropion-naltrexone. The incidence of hair loss was higher among the semaglutide group than the active comparator, bupropion-naltrexone. The adjusted hazard ratio for hair loss for all patients using semaglutide, for men, and for women compared to bupropion-naltrexone were 1.52 (95% confidence interval (CI):0.86-2.69), 0.86 (95% CI: 0.05-14.49), and 2.08 (95% CI: 1.17-3.72) respectively. Our results demonstrate an increased risk of hair loss with semaglutide in women. Future studies are required to ascertain the association between semaglutide and hair loss.

## INTRODUCTION

Glucagon-like peptide-1 (GLP-1) agonists are one of the most prescribed classes of drugs worldwide and are indicated for the treatment of diabetes or obesity. Approximately 1 in 8 people in the USA have used GLP-1 agonists^1^. Several studies have addressed the adverse event profiles of these drugs but have focussed mainly on the gastrointestinal adverse events^2^. Anecdotal data has alluded to hair loss as another adverse event associated with semaglutide. Data from the Food and Drug Administration (FDA) have reported more hair loss events with GLP-1 agonists compared to other antihyperglycemics^3^. However, to date, no epidemiologic study has examined this association in a real-life clinical setting especially in those taking semaglutide purely for weight loss.

## METHODS

Our data included a random sample of 16 million patients (2006-2020) from the IQVIA PharMetrics^®^ Plus for Academics. It captures 93% of all outpatient prescriptions and physician diagnoses in the US through the *International Classification of Diseases, Ninth and Tenth Revisions (ICD-9 or 10)*. We included new users of semaglutide, the most prescribed GLP-1 agonist, and the active comparator bupropion-naltrexone, a weight loss agent unrelated and chemically distinct to semaglutide. Those with a diagnosis of diabetes or use of an antihyperglycemic before cohort entry were excluded. This cohort was previously described by Sodhi^2^. Cohort members were followed from the first date of a study drug prescription to the first diagnosis of hair loss defined by the first ICD-9 (704.0X) or ICD-10 (L63.0, L63.1, L63.9, L64.0, L 64.8, L64.9, L65.0, L65.1, L 65.8, L65.9) code for the condition or termination of follow up. A Cox regression model was used to compute hazard ratios (HRs) that adjusted for age, sex, geographic location, depression, anemia, hypothyroidism, polycystic ovary syndrome, and use of oral corticosteroids. All analyses were performed using SAS version 9.4. Ethics approval was obtained by the University of British Columbia’s clinical research ethics board with a waiver of informed consent.

## RESULTS

Our cohort included 1,926 semaglutide users and 1,348 users of bupropion-naltrexone (Table 1). Bupropion-naltrexone users were younger than semaglutide users (Table 1). Incidence rates for hair loss for semaglutide and bupropion-naltrexone users was 26.5/1,000 and 11.8/1,000 person-years respectively (Table 1). The crude hazard ratio for hair loss for all patients taking semaglutide, for men, and for women was 1.30 (95% confidence interval (CI): 0.74-2.28), 0.70 (95% CI: 0.04-11.43), and 1.94 (95% CI: 1.09-3.54) respectively (Table 2). The adjusted hazard ratio for hair loss for all patients using semaglutide, for men, and for women compared to bupropion-naltrexone were 1.52 (95% CI: 0.86-2.69), 0.86 (95% CI: 0.05-14.49), and 2.08 (95% CI: 1.17-3.72) respectively (Table 2).

**Table 1.** Characteristics of semaglutide, liraglutide, and bupropion-naltrexone users.

|  | **Semaglutide** | **Bupropion-Naltrexone** |
| --- | --- | --- |
| **Number** | 1,926 | 1,348 |
| **Age (in years, mean ± STD)** | 54.8 ± 117 | 45.9 ± 11.3 |
| **Sex (%)** |  |  |
| **Male** | 46.7 | 18.6 |
| **Female** | 53.3 | 81.4 |
| **Median Follow-up in Years (interquartile range)** | 0.3-0.7 | 0.8-1.8 |
| **Incidence\*** | 26.5 | 11.8 |
| **Covariates (%):** |  |  |
| Depression | 20.5 | 14.1 |
| Hypothyroidism | 26.3 | 23.4 |
| Oral Steroids | 5.1 | 5.7 |
| Polycystic Ovary Syndrome | 5.1 | 5.7 |
| Anemia | 11.3 | 10.0 |
| **Region (in the USA):** |  |  |
| *Northeast* | 13.1 | 17.8 |
| *Southeast* | 32.0 | 32.2 |
| *Midwest* | 29.6 | 22.8 |
| *Southwest* | 11.4 | 14.2 |
| *West* | 13.6 | 12.8 |
| *Unknown* | 0.3 | 0.3 |

\*Incidence is rate per 1,000 person-years.

**Table 2.** Hazard ratios and 95% confidence intervals of hair loss among semaglutide users compared to bupropion-naltrexone users including an analysis stratified by sex.

|  | **Semaglutide** | **Bupropion-Naltrexone** |
| --- | --- | --- |
| **Total** |  |  |
| **Number of Events** | 23 | 27 |
| **Crude HR (95% CI)** | 1.30 (0.74-2.28) | 1 (Reference) |
| **Adjusted\* HR (95% CI)** | 1.52 (0.86-2.69) | 1 (Reference) |
| **Men** |  |  |
| **Number of Events** | 1 | 1 |
| **Crude HR (95% CI)** | 0.70 (0.04-11.43) | 1 (Reference) |
| **Adjusted\* HR (95% CI)** | 0.86 (0.05-14.49) | 1 (Reference) |
| **Women** |  |  |
| **Number of Events** | 22 | 26 |
| **Crude HR (95% CI)** | **1.94 (1.09-3.54)** | 1 (Reference) |
| **Adjusted\* HR (95% CI)** | **2.08 (1.17-3.72)** | 1 (Reference) |

\* Adjusted for covariates in table 1

## DISCUSSION

This study shows that semaglutide is associated with hair loss compared to users of bupropion-naltrexone. Although we found an increase in the overall HRs, the confidence intervals were imprecise leading to inconclusive results. However, when we stratified the results by sex, we note a greater than double increase in the HR in women.

Our results are in line with a clinical trial data for semaglutide (Wegovy^®^) that also showed an elevated risk of hair loss (3.3% in the Wegovy^®^ group compared to 1.4% in the placebo, RR=2.38, 95% 1.41-4.0) compared to placebo users^4^. Although not fully elucidated, it is postulated that hair loss secondary to semaglutide might be due to the physiological stress that rapid weight loss can induce, causing telogen effluvium and leading to increased hair shedding^5^. This may be more prominent with semaglutide as it is known to decrease weight more rapidly than bupropion-naltrexone, thus inducing greater physiological stress on the body and thus potentially greater hair loss^6^. Furthermore, in the Wegovy^®^ trial, those with greater than 20% body weight reduction reported alopecia more than those losing less than 20% of their body weight (5.3% versus 2.5%)^4^. This further lends credence that the mechanism for semaglutide induced hair loss may be due to the physiologic stress on the body that rapid weight loss induces, thus disrupting the hair cycle, leading to hair loss^3,4^. It is also possible that due to semaglutide’s appetite suppressive properties, patients have less food intake that can potentially cause nutrient deficiencies, and in particular, protein deficiencies which has been shown to be associated with hair loss^3^. Additionally, in conjunction with other potential adverse effects of semaglutide use such as nausea and gastroparesis^2^, total oral intake may decrease and loss of nutrients through vomitus can occur^3^.

Lastly, it has been hypothesized that semaglutide can lead to hormonal shifts that increase the risk of androgenic alopecia, whereby patients experience thinner hair and may potentially stop developing appreciable hair strands that can result in permeant hair loss, despite cessation of treatment^3,7^.

Limitations of our study include lack of data on semaglutide’s indication. Although we have excluded those with diabetes and those taking antihyperglycemics in the year prior to cohort entry, we cannot be certain that patients were taking semaglutide solely for weight loss.

Those considering using semaglutide strictly for weight loss might want to factor in hair loss as a possible limitation of these drugs, and in particular, women who may want to use semaglutide.

The risk benefit calculus to treatment initiation may be different for those with diabetes or morbid obesity and they may be more willing to accept hair loss as a potential risk of semaglutide treatment than those using semaglutide for recreational weight loss. Future studies are required to ascertain the association between GLP-1 agonists and hair loss.

## Data Availability

All data produced in the present study are available upon reasonable request to the authors.

## Author Contributions

Dr. Etminan had full access to all the data and takes responsibility for the integrity and accuracy of the data.

*Concept and design:* Etminan and Sodhi

*Acquisition, analysis, or interpretation of data:* All authors

*Drafting of the manuscript:* Etminan, Sodhi, Rezaeianzadeh, and Frey

*Critical revision of the manuscript for important intellectual content:* All authors

*Statistical analysis:* Kezouh

*Obtained funding:* Etminan

*Supervision:* Etminan

## Data Sharing Agreement

We will not be sharing the data, however we are open to answering any questions regarding collection, management, analysis, or interpretation of our data source.

